# The Value of a Regional ‘Living’ COVID-19 Registry and the Challenges of Keeping It Alive

**DOI:** 10.1101/2021.04.06.21255019

**Authors:** John Hanna, Tara Chen, Carlos Portales-Castillo, Donna Newhart, Katherine Schantz, Kathleen Rozzi, Jonathan Bress, Emil Lesho

## Abstract

**Background:** The need for rapid access to regularly updated patient data for hypothesis testing, surge planning, and epidemiologic investigations underscore the value of updated registries that clinicians, researchers, and policy makers can easily access for local and regional planning. We sought to create an adaptive, living registry containing detailed clinical and epidemiologic and outcome data from SARS-CoV-2-PCR-positive patients in our healthcare system.

**Methods:** From 03/13/202 onward, demographics, comorbidities, outpatient medications, along with 75 laboratory, 2 imaging, 19 therapeutic, and 4 outcome-related parameters were manually extracted from the electronic medical record of SARS-CoV-2 positive patients. These parameters were entered on a registry featuring calculation, graphing tools, pivot tables, and a macro programming language. Initially, two internal medicine residents populated the database, then professional data abstractors populated the registry. When the National Center for Immunization and Respiratory Diseases released their COVID-19 case report form for public access, we adapted it and used it on a browser-based, metadata-driven electronic data capture software platform. Statistics were performed in R and Minitab.

**Results:** At the time of this submission, 200,807 SARS-CoV-2 RT-PCR tests were performed on 107,604 distinct patients. 3699 (3.4%) of those have had positive results. Of those, 399 (11%) have had the more than 75 parameters full entered in the registry. The average follow-up period was 25 days (range 21-34 days). Age, male gender, diabetes, hypertension, cardiovascular disease, kidney disease, and cancer were associated with hospital admission (all p values < 0.01), but not ICU admission. Statin, ACEI-ARB, and acid suppressant use were associated with admission (all p values < 0.03). Obesity and history of autoimmune disease were not associated with need for admission. Supplemental oxygen, vasopressor requirement, and outpatient statin use were associated with increased mortality (all p values < 0.03).

**Conclusion:** A living COVID-19 registry represents a mechanism to facilitate optimal sharing of data between providers, consumers, health information networks, and health plans through technology-enabled, secure-access electronic health information. Our approach also involves a diversity of new roles in the field, such as using residents, staff, and the quality department, in addition to professional data extractors and the health informatics team.

However, due to the overwhelming number of infections that continues to accelerate, and the labor/time intense nature of the project, only 11% of all patients with COVID-19 had all parameters entered in the registry. Therefore, this report also offers lessons learned and discusses sustainability issues, should others wish to establish a registry. It also highlights the local and broader public health significance of the registry.

## Background

One dilemma in the attempt to deliver state-of-the-art therapeutics or understand emerging novel infectious pathogens is that the most current data is not always rapidly accessible to clinicians as soon as it becomes available. Hence, living systematic reviews have arisen as a potential solution to narrow the gap between evidence and practice.^1, 2^

Along the same lines, having live access with continuous updates of a patient data base using a standardized, well designed format and a user friendly, interactive software which allows us to also to more rapidly hypothesis test or identify patterns relevant to prevention motivated this effort. Additionally, data from one country or U.S. locality may not be generalizable to others, and data and experience from non-university or rural settings may be underrepresented in the medial literature, or not covered by media.^3^ Furthermore, rural areas are currently experiencing some of the biggest increases in new SARS-CoV-2 infections. Fourth, on June 19, 2020, the National Academies of Sciences, Engineering, and Medicine hosted a public meeting on “Data Needs to Monitor the Evolution of SARS-CoV-2”. Presenters agreed that regional surveillance nodes were needed. Last, the recently described “tragic data gap”, and the federal curtailment of reporting COVID-19 data to the national Health Safety Network, also provided motivation for this intervention.^4^ In the current climate, archiving detailed patient data in retrievable, easily analyzable, and share-friendly formats has become crucial for informing responses to current and future pandemics.^4^

Recent guidance^5^ and events^6, 7^ also underscore the added value having private, nongovernmental alternatives for collecting and analyzing large scale epidemiologic data.

We sought to implement an adaptive, ‘living’ registry capable of capturing detailed epidemiologic and clinical information from every patient diagnosed with SARS-CoV-2 infection, similar to and more granular than the Danish COVID-19 Cohort or TriNetX network. Examples of the living approach include the living rapid reviews in the Annals of Internal Medicine, and the living systematic review of the University of Toronto regarding secondary infections in patients with COVID-19. However, these are literature reviews and not registries.

Our ultimate goal is to have a minable source of detailed information updated on an ongoing basis, with minimal human effort, and linkable to SARS-CoV-2 whole genome sequences, and other registries across the United States.Perhaps, becoming one of many extra-governmental surveillance nodes in a regional or national network.

## Methods

The healthcare system consists of five acute care hospitals (ACH) and six long-term care facilities (LCTF) totaling 1056 ACH and 800 LCTF beds. It spans eleven suburban, rural and urban counties in the Finger-Lakes Region of NY. The first iteration of the registry was completed on 04/11/2020. Collection dates ranged from the first patient diagnosed w/ SAR-COV-2 on 03/13/2020 to the present.

In mid-March 2020, after the first case in our community, demographics, comorbidities, outpatient medications, along with 75 laboratory, 2 imaging, 19 therapeutic, and 4 outcome-related parameters were manually extracted from the electronic medical record of SARS-CoV-2 positive patients. These parameters were entered on a registry featuring calculation, graphing tools, pivot tables, and a macro programming language. Initially, two internal medicine residents populated the database, then professional data abstractors populated the registry.

All patients in the above healthcare system who had a positive PCR for SARS-CoV-2 from 03/13/2020 onward are included. At the time of this submission, 200,807 SARS-CoV-2 RT-PCR tests were performed on 107,604 distinct patients. 3699 (3.4%) of those have had positive results. Of those, 399 (11%) have had the more than 75 parameters full entered in the registry. The average follow-up period was 25 days (range 21-34 days).

When the National Center for Immunization and Respiratory Diseases released their COVID-19 case report form (Last updated May 5th 2020, available at https://www.cdc.gov/coronavirus/2019-ncov/php/reporting-pui.html for public access, we adapted it and used it on a browser-based, metadata-driven electronic data capture software platform.

Statistics were performed in R and Minitab.

## Results

Descriptive statistics are presented in Table 1, with mortality associations in Table 2. Age, male gender, diabetes, hypertension, cardiovascular disease, kidney disease, and cancer were associated with hospital admission, but not ICU admission. Statin, ACEI-ARB, and acid suppressant use were associated with admission as well. Obesity and history of autoimmune disease were not associated with need for admission. Supplemental oxygen and vasopressor and outpatient statin use were associated with increased mortality.

**Table 1:** Descriptive Statistics, Outpatient vs. Inpatient (ICU vs. non-ICU)

|  | Total (n=180) | Outpatient (n=92) | Inpatient (n=88) |  | p-value |
| --- | --- | --- | --- | --- | --- |
|  |  |  | ICU (n=35) | Non-ICU (n=53) |  |
| **Demographics** |  |  |  |  |  |
| Age | 55.0 (37.8, 68.0) | 45.5 (30.8,58.0)) | 62.5 (52.0, 73.0) |  | <0.0001 ** |
|  |  |  | 66.0 (55.0, 74.0) | 61.0 (50.0, 73.0) | 0.33 |
| Male Gender | 87 (48%) | 34/92 (37%) | 53/88 (60%) |  | 0.003 ** |
|  |  |  | 24 (69%) | 29 (55%) | 0.28 |
| Race | Black 40/163 (25%)<br>White 97/163 (60%)<br>Hispanic 18/163 (11%)<br>Asian 1/163 (1%)<br>Other 7/163 (4%) | 23/75 (31%)<br>42/75 (56%)<br>8/75 (11%)<br>0/75 (0%)<br>2/75 (3%) | 17/88 (19%)<br>55/88 (63%)<br>10/88 (11%)<br>1/88 (1%)<br>5/88 (6%) |  | 0.31 |
|  |  |  | 9 (26%)<br>23 (66%)<br>2 (6%)<br>1 (3%)<br>0 (0%) | 8 (15%)<br>32 (60%)<br>8 (15%)<br>0 (0%)<br>5 (9%) | 0.27 |
| BMI | 30.7 (26.3,37.8) | 30.9 (25.6,37.2) | 30.4 (27.2,38.3) |  | 0.43 |
|  |  |  | 31.2 (27.2, 38.6) | 30.0 (27.1, 37.9) | 0.78 |
| Current Smoker | 11/160 (7%) | 5/87 (6%) | 6/87 (7%) |  | 1.00 |
|  |  |  | 3 (9%) | 3/52 (6%) | 0.68 |
| Current or Former Smoker | 73/160 (46%) | 30/73 (41%) | 43/87 (49%) |  | 0.37 |
|  |  |  | 18 (51%) | 23/52 (44%) | 0.34 |
| Health Care Worker | 38/169 (22%)) | 27/81 (33%) | 11/88 (13%) |  | 0.002 ** |
|  |  |  | 4 (11%) | 7 (13%) | 1.00 |
| **Comorbidities** |  |  |  |  |  |
| Morbid Obesity | 24/179 (13%) | 8/91 (9%) | 16 (18%) |  | 0.10 |
|  |  |  | 6 (17%) | 10 (19%) | 1.00 |
| Chronic Resp Dz | 36/179 (20%) | 13/91 (14%) | 23 (26%) |  | 0.07 |
|  |  |  | 7 (20%) | 16 (30%) | 0.41 |
| CKD/ESRD | 20/179 (11%) | 6/81 (7%) | 14 (16%) |  | 0.06 |
|  |  |  | 9 (26%) | 5 (9%) | 0.07 |
| HTN | 83/179 (46%) | 27/91 (30%) | 56 (64%) |  | <0.0001 ** |
|  |  |  | 23 (66%) | 33 (62%) | 0.92 |
| DM | 49/179 (27%) | 15/91 (16%) | 34 (39%) |  | 0.002 ** |
|  |  |  | 19 (54%) | 15 (28%) | 0.08 |
| Cardiovascular Dx | 44/179 (25%) | 11/91 (12%) | 33 (38%) |  | 0.0002 ** |
|  |  |  | 14 (40%) | 19 (36%) | 0.43 |
| Cancer Dx | 6/179 (3%) | 0/91 (0%) | 6/88 (7%) |  | 0.01 * |
|  |  |  | 4 (11%) | 2 (4%) | 0.21 |
| Autoimmune Dx | 4/179 (2%) | 2/91 (2%) | 2 (2%) |  | 1.00 |
|  |  |  | 2 (6%) | 0 (0%) | 0.16 |
| Liver disease Dx | 4/179 (2%) | 0/91 (0%) | 4/88 (5%) |  | 0.06 |
|  |  |  | 3 (9%) | 1 (2%) | 0.30 |
| **Medications** |  |  |  |  |  |
| ACE/ARB use | 39/179 (22%) | 12 (13%) | 27/87 (31%) |  | 0.006 ** |
|  |  |  | 15/34 (44%) | 12 (23%) | 0.06 |
| Acid suppression medication use | 61/179 (34%) | 23 (25%) | 38/87 (44%) |  | 0.01 * |
|  |  |  | 18/34 (53%) | 20 (38%) | 0.24 |
| Statin use | 55/179 (31%) | 16 (18%) | 39/87 (45%) |  | 0.0001 ** |
|  |  |  | 18/34 (53%) | 21 (40%) | 0.32 |
| Sick Contact |  |  |  |  |  |
| COVID Contact | 44/166 (27%) | 22/80 (28%) | 22/86 (26%) |  | 0.92 |
|  |  |  | 4/34 (12%) | 18/52 (35%) | 0.03 * |
| Sick Contact | 86/159 (54%) | 42/74 (57%) | 44/85 (52%) |  | 0.63 |
|  |  |  | 16 (46%) | 28/50 (56%) | 0.48 |
| Time from Symptoms to first presentation(days) | 5.0 (3.0, 9.5) | 5.0 (3.0, 9.0) | 5.0 (3.0, 9.3) |  | 0.91 |
|  |  |  | 4.0 (2.0, 7.0) | 6.0 (3.0, 10.0) | 0.04 * |
| Labs |  |  |  |  |  |
| WBC | 6.0 (4.7, 7.8) | 5.9 (4.5,7.8) | 6.1 (4.8,7.9) |  | 0.28 |
|  |  |  | 6.8 (4.9, 7.7) | 5.9 (4.7,8.0) | 0.55 |
| Lymphocytes | 1.0 (0.7, 1.4) | 1.1 (0.8, 1.5) | 1.0 (0.6, 1.3) |  | 0.18 |
|  |  |  | 0.9 (0.6, 1.4) | 1.1 (0.7, 1.3) | 0.68 |
| ANC | 4.1 (3.1, 6.0) | 3.7 (2.6,4.7) | 4.2 (3.5, 6.3) |  | 0.08 |
|  |  |  | 4.9 (3.7, 6.5) | 4.1 (3.2, 5.8) | 0.33 |
| D-Dimer | 826 (601, 1418) | Only 2 values | 889 (634, 1439) |  | XXXXXX |
|  |  |  | 831 (642,1349) | 946 (631, 1450) | 1.00 |
| LDH | 302 (261, 399) | Only 1 value | 336 (266, 399) |  | XXXXXX |
|  |  |  | 362 (323,462) | 293 (254, 389) | 0.11 |
| Procalcitonin | 0.12 (0.06, 0.19) | No values | 0.12 (0.06, 0.19) |  | XXXXXX |
|  |  |  | 0.10 (0.06, 0.21) | 0.12 (0.07, 0.19) | 0.62 |
| CRP | 51 (28, 119) | Only 1 value | 52.5 (29.2, 119.2) |  | XXXXXX |
|  |  |  | 121 (44,135) | 51 (22,102) | 0.07 |
| CK | 97 (69,204) | No values | 97 (69, 204) |  | XXXXXX |
|  |  |  | 71 (60, 110) | 112 (74, 204) | 0.31 |
| Imaging |  |  |  |  |  |
| Presentation CXR positive | 59/112 (53%) | 7/26 (27%) | 52/86 (60%) |  | 0.005 ** |
|  |  |  | 22 (63%) | 30/51 (59%) | 0.88 |
| Presentation CT positive | 19/22 (86%) | 0/3 (0%) | 19/19 (100%) |  | 0.0006 ** |
|  |  |  | 8/8 (100%) | 11/11 (100%) | 1.00 |

**Table 2:** Chi Square Test for Associations with Morality.

| Medication | No Died | Yes Died | No D/C | Yes D/C | P value |
| --- | --- | --- | --- | --- | --- |
| Hydroxychloroquine | 13 | 10 | 74 | 35 | 0.296 |
| Kaletra | 22 | 2 | 105 | 4 | 0.319 |
| Anti IL-6 | 23 | 1 | 108 | 1 | 0.236 |
| Pressor Support | 13 | 11 | 100 | 9 | 0.001 |
| Non- inhaled Steroids | 17 | 7 | 91 | 17 | 0.123 |
| Supplemental O2 | 5 | 19 | 48 | 60 | 0.033 |
| HFNC | 18 | 6 | 96 | 12 | 0.073 |
| Vit D Home | 11 | 8 | 74 | 33 | 0.334 |
| Statin Home | 6 | 14 | 66 | 42 | 0.010 |

|  | No Died | No D/C | ACE Died | ACE D/C | ARB Died | ARB D/C | Both Died | Both D/C | P value |
| --- | --- | --- | --- | --- | --- | --- | --- | --- | --- |
| ACE/ARB | 10 | 73 | 3 | 22 | 4 | 12 | 0 | 6 | 0.391 |

|  | No Died | No D/C | PPI Died | PPI D/C | H2B Died | H2B D/C | P Value |
| --- | --- | --- | --- | --- | --- | --- | --- |
| Home Suppressor | 7 | 66 | 11 | 31 | 1 | 6 | 0.061 |

Factors associated with morality from 399 COVID-19 patents in Western New York from March 15, 2020 to April 15, 2020

## Discussion

The registry revealed local patterns not apparent from less granular databased such as the CDC or county/state health departments, or from reports of other populations. For example, the average length of stay (LOS) for all admitted patients was 8 days (SD 8.6). The average LOS for patients who did and did not require ICU-level care was 13.5 (SD 10.4) and 5.9 (SD 6.8), respectively. This is significant because remdesivir was not available during the follow up period for this initial cohort, and the LOS without it was already shorter than the 15-day outcome measure used in the final report.^8^ Another example is a meta-analysis that found 5 studies, ACEI or ARB use was associated with a lower risk for severe illness. According to the authors, those results “do not provide enough evidence to draw conclusions about the potential efficacy of these medications in treating COVID-19”.^2^ In contrast, we found use of angiotensin inhibitors and angiotensin receptor blockers was higher in admitted vs. ED treated-and-released patients (p ≤ 0.001). A third example involves a Chinese report which showed that statin treatment was associated with lower mortality.^9^ However in our population, statin use was higher in admitted patients compared to patients who did not require admission (all p ≤ 0.001). Statin use was also higher in those who died than those who survived (Table 2) Last, in New York City, viral load was correlated with risk of intubation, but in our more rural and suburban area, we observed that viral load did not correlate with severity of illness.^10, 11^

Registry challenges, pitfalls, and threats to sustainability are presented in Table 3. Manual data extraction into the original spreadsheet became prohibitively labor intensive and analytically unmanageable as the number of new cases rapidly increased. Real-time abstraction needed to be suspended for several weeks while contents were transferred to the metadata-driven electronic data capture software platform. As of 06/14/2020, registry personnel were able to manually populate all the parameters mentioned above from 11% (399) of the patients who tested positive (236 ambulatory, 163 inpatient). Mean follow up period for that group for the descriptive statistics presented below 25 days (range 21-34 days).

**Table 3:**
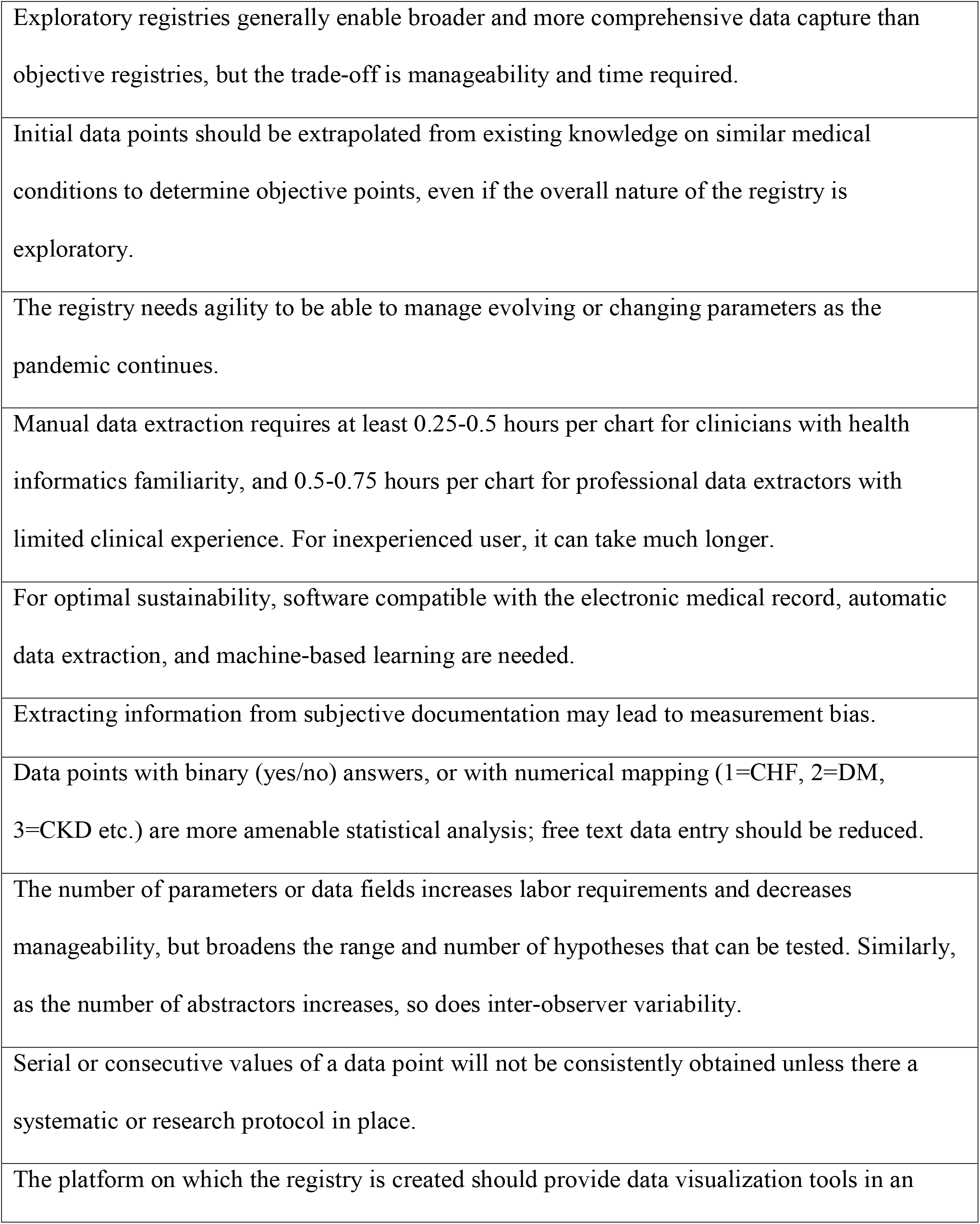
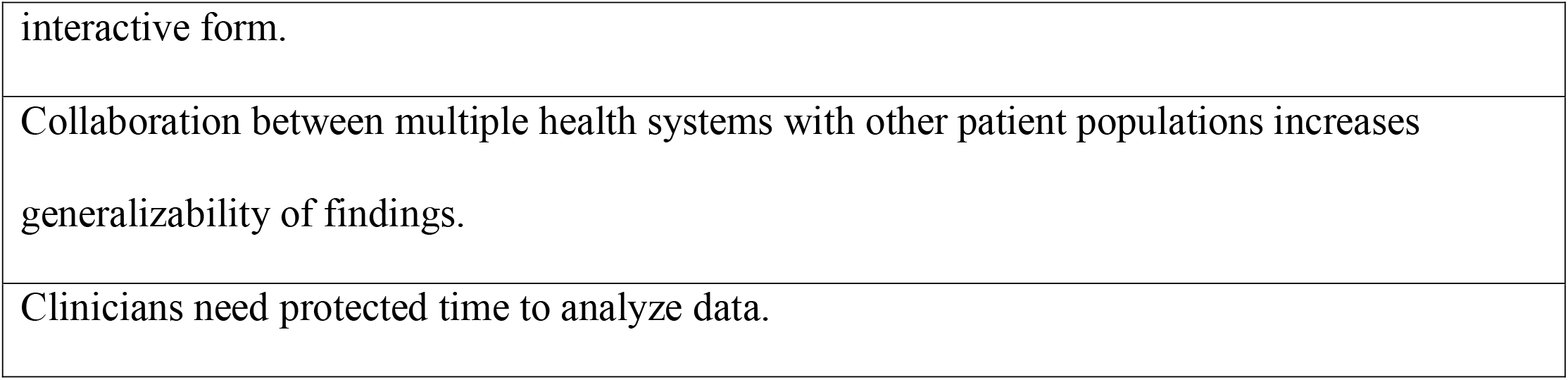
Considerations and Lessons Learned for Creating Registries.

For diseases with lower incidence rates, the sustainability of the initial approach would have been be achievable, but the approach became unsustainable given epidemic/pandemic-level volumes of new cases daily and weekly. For optimal sustainability, 2 full time experienced data extractors, automation and machine-based learning would be required.

## Conclusion

A living COVID-19 registry represents a mechanism to facilitate optimal sharing of data between providers, consumers, health information networks, and health plans through technology-enabled, secure-access electronic health information. Our approach also involves a diversity of new roles in the field such as using residents, staff, and the quality department, in addition to professional data extractors and the health informatics team.

However, due to the overwhelming number of infections that continues to accelerate, and the labor/time intense nature of the project, only 11% of all patients with COVID-19 had all parameters entered in the registry. Therefore, this report also offers lessons learned and discusses sustainability issues, should others wish to establish a registry. It also highlights the registry’s local and broader public health significance.

The registry is well suited to address a range of hypothesis and is being leveraged by other researchers in our system. It provides a resource for researchers, policy makers, surge planning, and ultimately linking detailed clinical-demographic data to whole genome sequencing data for precision epidemiology. This approach has been successfully used to improve outcomes for drug-resistant bacteria.^12^ Last, once established, the informatics algorithms and parameters can be applied to patients infected with any high-consequence or novel pathogen.

## Data Availability

NA

